# Dynamical balance between the transmission, intervention of COVID-19 and economic development

**DOI:** 10.1101/2020.08.13.20174755

**Authors:** Zhaowang Zhang, Hualiang Lin, Guanghu Zhu

## Abstract

The current explosive outbreak of coronavirus (COVID-19) is posing serious threats to public health and economy around the world. To clarify the coupling mechanism between this disease and economy, a new dynamical system is established. It is theoretically proved that the basic reproduction number is a nonlinear combination of parameters regarding disease transmission, intervention and economy effect, which totally determines the stability of the disease-free and endemic equilibria. Further results indicate the existence of interaction and mutual restraint among the transmission, intervention and economy, in which strong coupling of COVID-19 and economy would trigger disease outbreak and form poverty trap, while adaptive isolation of at-risk population could effectively reduce morbidity at the cost of least economic loss. Our findings can offer new insights to improve the intervention strategies against COVID-19.

## 1. Introduction

In late December 2019, a novel coronavirus disease (COVID-19) first emerged in Wuhan, China, and rapidly spread to other countries [1]. As of July 31, 2020, 216 countries are hit by COVID-19 pandemic with over 16 million confirmed cases and about 0.65 million deaths. WHO claimed the pandemic is still accelerating, and its various impacts may last for decades [2].

A series of unprecedented intervention measures have been taken to control the outbreak of COVID-19. Non-pharmaceutical interventions mainly focus on social distancing, including travel restrictions, quarantine of patients, closure of schools and workplaces, suspension of public transportation, and wearing masks. Such strategies are playing significant roles in reducing the morbidity and mortality [3], and at the same time, they directly/indirectly hinder economic development [4]. For example, Hubei’s GDP shrank by 39.2% in the first quarter compared with the previous one. It is estimated that the reduction in GDP range from 4% under alert level 1 up to 37% under level 4 in New Zealand [5]. Hence, in the face of COVID-19, it is of great significance to balance the relationship between economy and public health, which can help more effectively mitigate the disease burden with less economic loss.

Mathematical models are widely used for clarifying the transmission patterns of infectious diseases. They are particularly employed to illustrate the interplay between economic growth and epidemic ecology [6, 7]. For example, by analyzing coupled model of economic growth and disease transmission, two possible mechanisms were given to escape the poverty trap [6], and a significant empirical link was found between tuberculosis and economic production systems [7]. Since the occurrence of COVID-19, many mathematical models were established aiming at predicting propagation trends [8], tracing transmission patterns [9], estimating infection risk [10], and evaluating intervention effects [11]. However, recent studies on the relation between COVID-19 and economy only stays in descriptive analysis, and little work is found on the coupling mechanism between them.

To fill the knowledge gap, a new coupled model of COVID-19 transmission and economic growth is established by ordinary differential equations. The dynamical behaviors of the model are explored by using stability theory and numerical analysis, and particularly the dynamical linking between disease transmission/control and economic development is inferred.

## 2. Model formulation

Based on the compartmental principle and COVID-19 epidemiology, human population is divided into five parts which are susceptible, latent, infectious, cured and recovered. The GDP per capita and the proportion of population at the corresponding states at time *t* are represented by *g*(*t*), *s*(*t*), *l*(*t*), *i*(*t*), *c*(*t*) and *r*(*t*), respectively.

The specific transmission process is as follows: susceptible individuals can be infected by contacting latent and infectious individuals, and then go through an incubation period 1/*η*. They receive treatment after an infected period *1*/*δ* and finally recover. It is assumed that the total density of human is constant as unit, in which birth rate equals to death rate *b*. For disease control, parts of susceptible, latent and infectious people are isolated, the proportions of which are denoted by *q*_1_, *q*_2_, and *q*_3_, respectively. The corresponding transfer flowchart of the model is shown in Figure 1.

**Figure 1:**
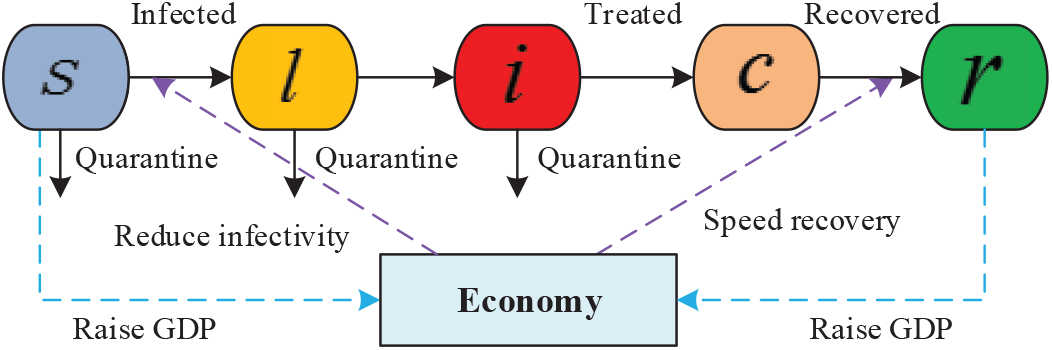
Flow diagram on the dynamical interactions between COVID-19 transmission and economic development.

The dynamics of COVID-19 and economy are inexorably linked. First, the working environment and living conditions will be improved through economic growth, which can further reduce the morbidity and mortality of COVID-19. Second, economic growth will make the treatment measures more standardized and perfect, which can further shorten the time of treatment and recovery. The economic growth model here is modelled using the Logistic equation [12], where COVID-19 patients and quarantine people could not work, so GDP growth will be constrained by COVID-19 epidemic. Based on the above description, the dynamical model was written by the following differential equations:

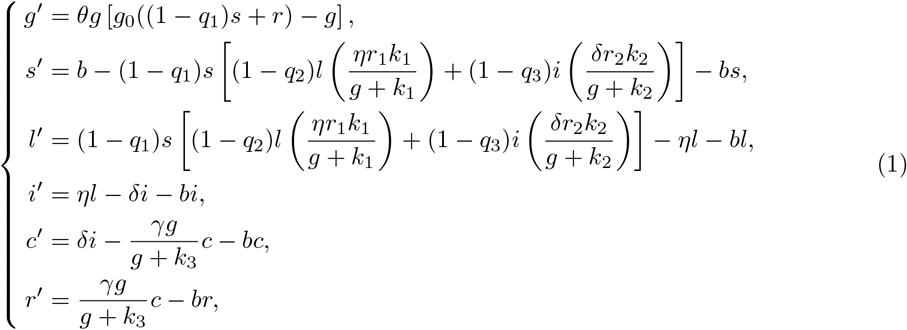

where the parameter *θ* and *g*_0_ represent the growth rate of GDP per capita, and the maximum income in absence of disease; *r*_1_ (*r*_2_) is the original reproduction number contributed by individuals in incubation (infectious) states, which equals the product of effective contact rates and time duration staying in the corresponding state. Here the infectivity and recovery rates are nonlinear functions of GDP per capita [12], with *k_i_* as the weighting coefficient.

## 3. Basic reproduction number and stability

Direct computation yields that the disease-free equilibria of system (1) include *e*_1_ = (0, 1, 0, 0, 0, 0) and *e*_2_ = (*g*_0_(1 − q_1_), 1, 0, 0,0,0). It is easy to prove that *e*_1_ is unstable, i.e. GDP can not be zero without COVID-19 epidemic. Using the next generation matrix, the basic reproduction number of system (1) is calculated as

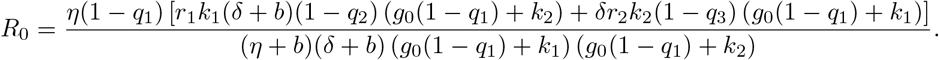

### Theorem 3.1

When *R*_0_ < 1, the disease-free equilibrium *e*_2_ of system (1) is locally asymptotically stable.

#### Proof

The Jacobi matrix of system (1) at *e*_2_ is

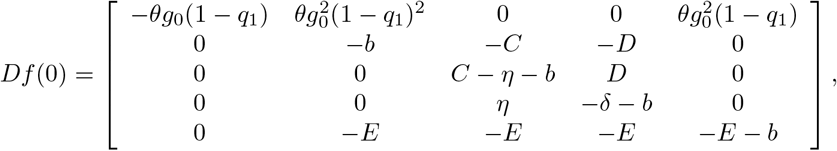

where

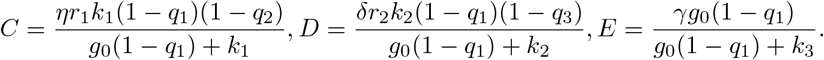

The characteristic equation of *Df* (0) is

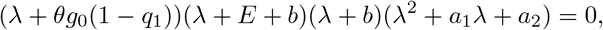

where *a*_1_ = *δ* + *b* + *η* + *b* − *C* and *a*_2_ = (*η* + *b* − *C*)(*δ* + *b*) − *D* = (*δ* + *b*)(*η* + *b*)(1 − *R*_0_). Since the eigenvalues *λ*_1_ = −*θg*_0_(1 − *q*_1_), *λ*_2_ = −*E* − *b* and *λ*_3_ = −*b* are negative, it only needs to consider the solution of *F*(*λ*) = *λ*^2^ + *a*_1_*λ* + *a*_2_ =0. When *R*_0_ < 1, it follows that *a*_1_ > 0 and *a*_2_ > 0. According to Hurwitz criterion, *F*(*λ*) has two eigenvalues of negative real part. Hence *e*_2_ is locally asymptotically stable when *R*_0_ *<* 1.

### Theorem 3.2

When *R*_0_ < 1, the disease-free equilibrium e_2_ of system (1) is globally asymptotically stable.

#### Proof

A Lyapunov function is defined as

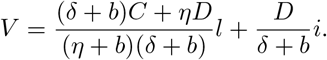

For *R*_0_ < 1, the derivatives of *V* along model (1) is

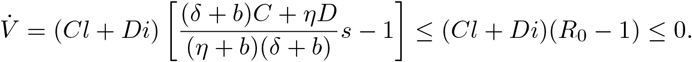

Since 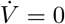 only if *l* = *i* = 0 or *R*_0_ = 1, and the maximum invariant set in 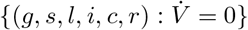 is the singleton {*e*_2_}, it follows from LaSalles invariance principle that *e*_2_ is globally stable.

### Theorem 3.3

When *R*_0_ > 1, then there exists an endemic equilibrium *P*\* in system (1), which is locally asymptotically stable.

#### Proof

Let the right side of system (1) be zero. It is obtained that

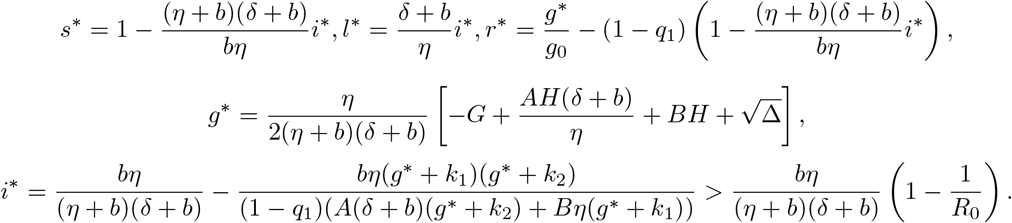

where

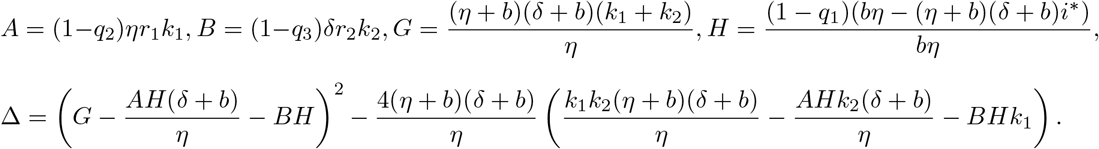

For *i*^*^ ≠ 0, when *R*_0_ > 1, let

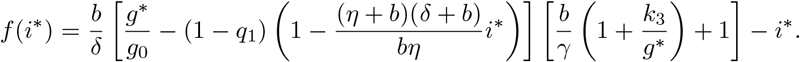

Direct calculation yields that f (0) < 0, *f* (1) > 0. It follows that when *R*_0_ > 1 there exists an endemic equilibrium *P*\*.

The Jacobi matrix of the system at *P\** is

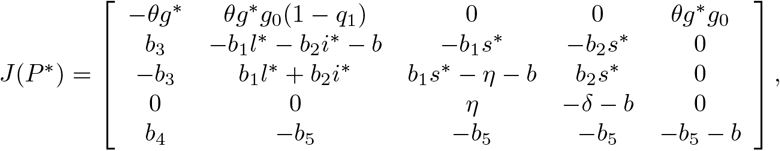

where

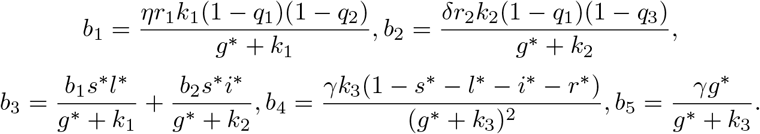

The characteristic equation of *J*(*P*\*) is *λ*^5^ + *m*_1_*λ*^4^ + *m*_2_*λ*^3^ + *m*_3_*λ*^2^ + *m*_4_*λ* + *m*_5_ = 0, where

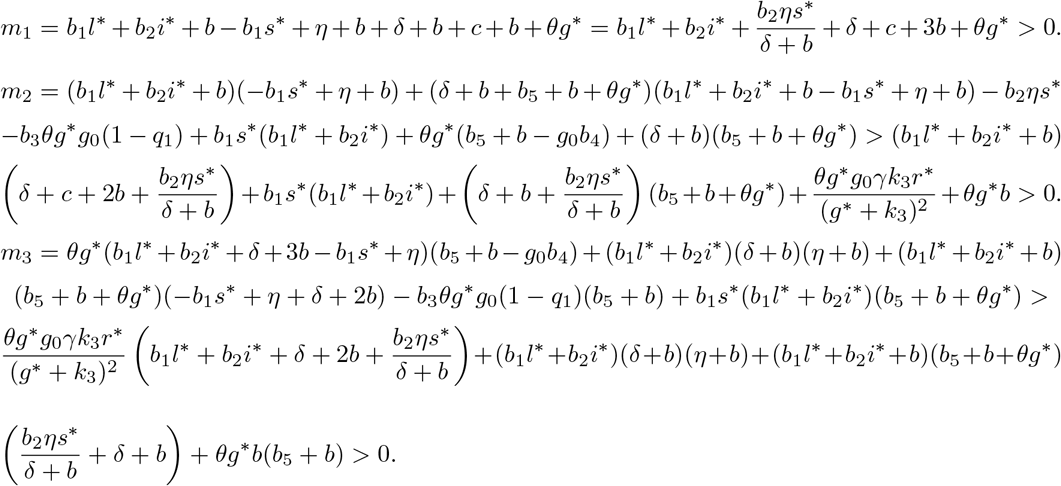

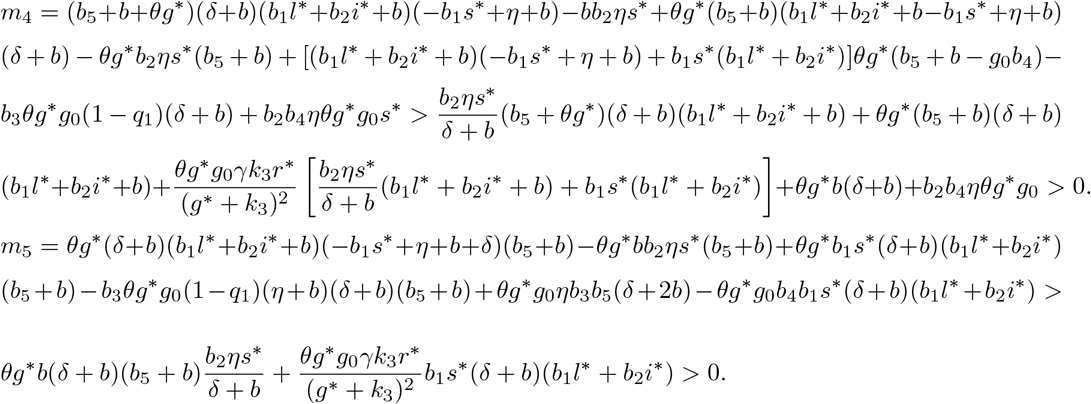

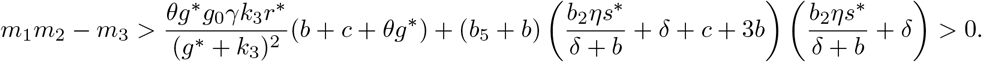

Direct computation yields that Δ_3_ > 0, Δ_4_ > 0. According to Routh-Hurwitz criterion, the endemic equilibrium *P*\*is locally asymptotically stable.

## 4. Numerical analysis

The model framework is activated by concrete parameters with biological background, which allows for further digging the dynamical interactions between COVID-19 and economic growth. The time unit of the parameters is per month or month. According to data of China’s National Bureau of Statistics in 2019, the GDP per capita (*g*_0_) and the birth/death rate are respectively adopted to be 5908 RMB and 0.0011 (with life expectancy as 76.34 years). It is estimated from the China’s GDP curve that the coefficient *θ* is 4.40 × 10^−4^. The basic reproduction numbers contributed by individuals in incubation and infected states are specified as *r*_1_ = 0.4 and *r*_2_ = 2.68 [8]. The mean incubation period 1/*η* is 1/6 [13]. Based on the epidemiology survey, the time span from illness onset to be treated and the shortest period of treatment are estimated to be 1/*δ* = 0.2 and 1/*γ* = 1/3, respectively. In simulations, the proportions of humans in different states being quarantined and the weighting coefficient of economic effect are variable. It is imposed that the initial values of GDP per capita equals the positive equilibrium in case of no infection.

Figure 2 shows the effects of different coupling weight coefficient on the evolutions of COVID-19 and economy. It is found that when infectivity weight of GDP (*k*_1_ and *k*_2_) increase from 0 to 6000, the basic reproduction number *R*_0_ will increase from 0.10 to 1.55. That is, large coupling weights between the infectivity and economy may easily trigger the outbreak of COVID-19, which would formulate a poverty trap. It is further observed that (1) when *R*_0_ *<* 1, the incidence declines directly and rapidly, and GDP per capita increases slowly until stabilizes; (2) When *R*_0_ > 1, followed by a sharp rise, the incidence quickly reach an peak and then keep stable in very low level. At the same time, the GDP per capita falls rapidly and increase gradually to a stable level. It is clear that the outbreak of COVID-19 causes damage to economic growth, and it will take over ten years for economy to recover. It should be noted that the treatment weight coefficient of GDP (*k*_3_) could not work on *R*_0_ and the incidence. Yet large *k*_3_ means that the treatment cycle is much longer, which would leads to lower level of economic growth.

**Figure 2:**
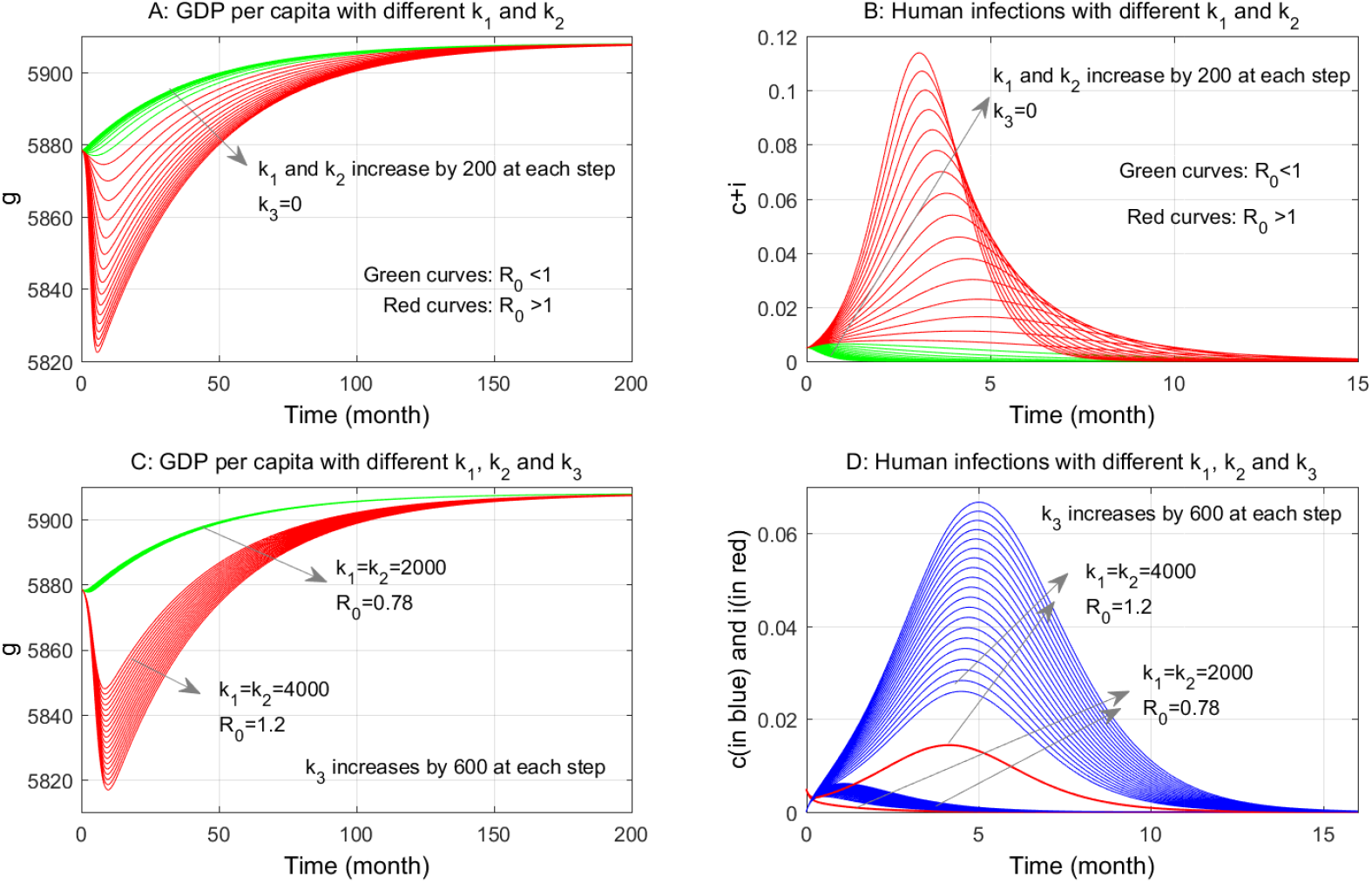
Time evolutions of GDP per capita and human infections with different economic effect weights *k*_1_, *k*_2_ and *k*_3_. The isolation rates are *q*_1_ = *q*_2_ = *q*_3_ = 0.

Figure 3 displays the restrictive relationships of COVID-19 and economy in case of implementing quarantine measures with different intensity. It is found that both of constant and adaptive isolation can work on COVID-19 control and economic situation, in which some interesting patterns are observed: (1) Adaptive isolation could be more effective in decreasing morbidity, but it also prolongs the epidemic in very low prevalence rate; (2) Isolating susceptible individuals would greatly damage economic growth and could slightly release infection burden; (3) Adaptively isolating the people in latent and infected states not only can effectively reduce incidence rate, but also minimize the impacts on economy. Particularly, if without quarantine, the economy would get hit due to high morbidity and less labor, which would suffer much heavier blow compared with implementing adaptive isolation.

**Figure 3:**
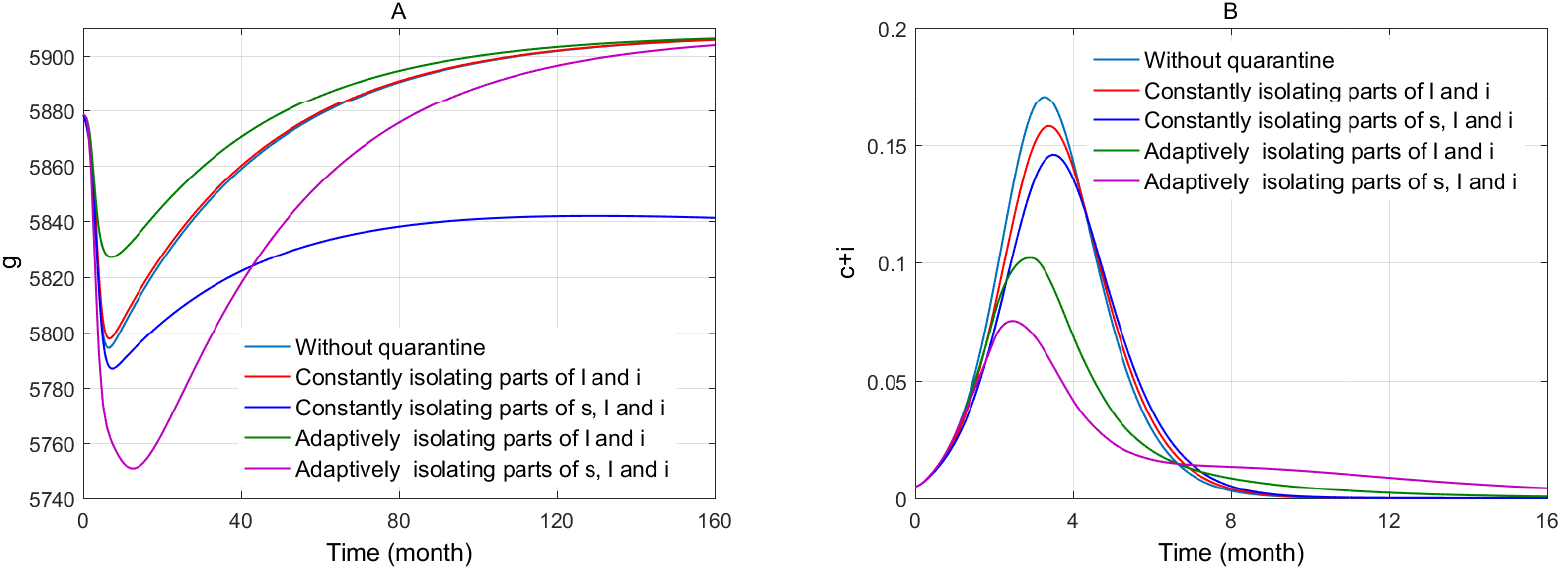
Time evolutions of GDP per capita and human infections with different intervention strategies, where *k*_1_ = *k*_2_ = *k*_3_ = 6000. The isolation rate in constant (adaptive) isolation at time *t* is defined as constant *q* (4 times of *c* at time *t* − 1), with equal averages for comparability.

Finally, sensitivity index of *R*_0_ to model parameters is calculated by using formula 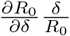 (for parameters *δ*) [14]. As show in Table 1, it is found that the original reproduction numbers (*r_i_*), and the weighting coefficients of economy on infectivity (*k_i_*) are positively correlated with *R*_0_, indicating that releasing the coupling weight between COVID-19 and economy, and especially cutting down contacts with infected people can effectively lower infection risk. Furthermore, the growth rate of GDP per capita (*θ*), and the maximum income in absence of disease (go), as well as the isolation rates (*g*_0_) are negatively associated with *R*_0_, indicating that economic development and quarantine strategies can significantly reduce infection risk.

**Table 1:** Sensitivity indices (SI) of R0 to model parameters.

| Parameter | $r_2$ | $g_0$ | $k_2$ | $q_3$ | $q_1$ | $r_1$ | $k_1$ | $q_2$ | $b$ | $\delta$ | $\eta$ |
| --- | --- | --- | --- | --- | --- | --- | --- | --- | --- | --- | --- |
| SI | 0.87 | -0.44 | 0.38 | -0.22 | -0.14 | 0.13 | 0.06 | -0.03 | -0.00038 | 0.00019 | 0.00018 |

## 5. Discussion

In this paper, a new dynamical model is developed for coupling the transmission, intervention of COVID-19 and economic growth. Mathematical analysis verifies that the basic reproduction number *R*_0_ is the sharp threshold to determine the outbreak of the disease. The value of *R*_0_ greatly depends on intrinsic transmissibility (e.g., infection rate, incubation period, and infected period), coupling weight, and intervention strategies (e.g., early detection and treatment, quarantine). The results indicate that there is a mutual restraining mechanism between COVID-19 transmission and economic growth. Strong impact of economy on COVID-19 infection would trigger disease outbreak and raise morbidity, and it also hold back economic progress, resulting in a poverty trap. In this case, providing financial assistance for attacking areas or patients can release the connection between infection and economy loss, which could help to escape the poverty trap stemmed from COVID-19 disease. Furthermore, implementing quarantine measures can help to contain the propagation of COVID-19, but its impact on economy is inconsistent. Isolating healthy people would damage economy. While adaptively isolating those people in latent and infected states as many as possible is best for both sides, which can greatly reduce human infection and yields a minimal impact on economic development. That requires timely detection and isolation of those persons at risk (e.g., the people who once contact with patient or has been to infected regions), and guarantees that healthy persons can work in safe environment. The results can provide scientific theoretical reference for COVID-19 epidemic control and policy formulation.

## Data Availability

The data is availabe from the corresponding author, upon reasonable request.

